# Sleep-Derived Features From Multi-Night In-ear EEG Identify Patterns Linked To Mild Cognitive Impairment

**DOI:** 10.1101/2025.11.11.25339999

**Authors:** Yashar Kiarashi, Emily L. Giannotto, Mohsen Motie-Shirazi, Amy D. Rodriguez, Allan I. Levey, Gari D. Clifford

**Affiliations:** Department of Biomedical Informatics, School of Medicine, Emory University, Atlanta, GA, 30322, USA; Department of Neurology, School of Medicine, Emory University, Atlanta, GA, 30322, USA; Atlanta VA Medical Center, Center for Visual and Neurocognitive Rehabilitation, Decatur, GA 30033, USA; School of Electrical and Computer Engineering, Georgia Institute of Technology, Atlanta, GA, 30332, USA

**Keywords:** Mild cognitive impairment, Sleep, In-ear EEG, AI, Wearable

## Abstract

**INTRODUCTION:** We investigated whether sleep features from multi-night, at-home in-ear EEG could distinguish mild cognitive impairment (MCI) from cognitively normal (CN) older adults, independent of demographic confounds.

**METHODS:** Forty-three older adults (24 MCI, 19 CN) completed 361 overnight in-ear EEG recordings at home (4.44 ± 2.29 nights per participant). Deep learning-derived embeddings were compared to bandpower features. We controlled for age and sex to identify sleep-specific features of MCI beyond demography.

**RESULTS:** Deep embeddings outperformed bandpower features (F1: 0.76 vs. 0.67) in classifying MCI from CN. Multi-night aggregation improved AUROC from 0.61 to 0.77, reduced within-subject variability 1.6-fold, and enhanced group separability based on Cohen’s d improving from 0.65 to 0.96. While some features correlated with age, MCI discrimination remained significant after controlling for demographics.

**DISCUSSION:** Multi-night in-ear EEG captures sleep features that reliably distinguish MCI from CN individuals beyond age-related effects, offering a scalable approach for at-home sleep monitoring.

## 1 INTRODUCTION

Sleep disturbances represent one of the earliest and most consistent changes associated with cognitive decline, emerging years before clinical symptoms of mild cognitive impairment (MCI) and Alzheimer’s disease (AD) manifest ^[1;2;3;4;5;6]^. These disruptions likely reflect the earliest pathophysiological changes in AD, as accumulating amyloid-*β* and tau proteins disrupt sleep-wake regulatory circuits in the brain, while impaired glymphatic clearance during sleep creates a feed-forward cycle where poor sleep quality accelerates protein aggregation ^[7;8]^.

Recent advances in wearable electroencephalography (EEG) technology ^[9;10;11;12;13]^and digital biomarker development have positioned sleep monitoring as a potentially transformative tool for early detection and monitoring of neurodegenerative diseases. Among wearable technologies, in-ear EEG^[14]^ offers unique advantages for longitudinal monitoring, providing clinical-grade signal quality while maintaining user comfort and compliance over extended periods. Unlike traditional polysomnography (PSG) or scalp-based devices, in-ear systems enable unobtrusive data collection that preserves natural sleep patterns while capturing the subtle EEG features necessary for sensitive and accurate detection of sleep microarchitecture. The convergence of sleep medicine, digital health, and aging research offers a unique opportunity to develop accessible with the objective on finding distinct changes in sleep patterns which are associated with cognitive disorders. Importantly, prior work by Lucey et. al, as one of the leading work in using wearables for sleep monitoring at home, has demonstrated that specific features of non-REM and slow-wave sleep are closely linked to amyloid and tau deposition, supporting the potential of EEG-based sleep metrics as early indicators of Alzheimer’s pathology ^[15;16]^. Their findings highlight that alterations in slow-wave sleep, spindles, and sleep architecture not only occur prior to measurable cognitive symptoms but also predict progression from normal cognition to MCI, further motivating longitudinal EEG monitoring approaches.

Most sleep studies in aging populations still rely on single night recordings ^[17;18]^ or actigraphy ^[19;20]^. This approaches obscures the night-to-night variability now recognized as a hallmark of prodromal neurodegeneration. Emerging evidence suggests that fluctuations in sleep-driven features across consecutive nights track synaptic homeostasis and glymphatic clearance, key processes that become dysregulated early in neurodegenerative disease progression. Specifically, alterations in slow-wave sleep characteristics, REM sleep fragmentation, sleep spindle density, and circadian rhythm stability have emerged as promising digital biomarkers for detecting prodromal neurodegeneration. Capturing these dynamics demands longitudinal monitoring capable of preserving the temporal structure of sleep while accommodating participants in their natural home environment.

In this study, we collected and used a novel dataset comprising seven consecutive nights of sleep data collected via in-ear EEG from older adults with MCI and their cognitively normal care partners. This unique dataset enables investigation of within-night sleep microarchitectural patterns while averaging out night-to-night noise, addressing a critical gap in sleep research where most studies rely on single-night recordings that are confounded by transient variability unrelated to disease.

## 2 METHODS

### 2.1 Participants

Participants for this study were recruited from the Charlie and Harriet Shaffer Cognitive Empowerment Program (CEP) at Emory University. The CEP is a six-month research program for persons with diagnosed MCI and their care partners, provides therapeutic education and programs across the following domains: physical activity, emotional wellbeing, nutrition, cognitive strategies, and home and personal safety. 67 CEP participants with MCI received are required to have a clinical diagnosis from the Emory Cognitive Neurology Clinic or the Goizueta Alzheimer’s Disease Research Center. Additional requirements include speaking English, being able to participate in group activities without severe behavioral disturbance, and ambulate and toilet independently. All CEP participants with MCI were eligible to enroll in the present study. Care partners were only eligible to enroll in the study if their care recipient also enrolled in the study, they shared a residence, and they agreed to participate together. Participants were enrolled as MCI-care partner dyads whenever possible.

Consent was obtained from 55 participants (*N* = 32 MCI, 22 CN) under Emory University approved IRB#: 00003011. Overnight sleep EEG recordings were obtained from 43 older adults using the NextSense in-ear EEG device (Figure 1 1 a,b). Three participants with MCI and one care partner withdrew from the study before sleep data collection began and four participants with MCI and 2 care partners only partially completed the study. In addition, two participants with MCI were unable to wear the in-ear EEG device due to ear anatomy.

**FIGURE 1.**
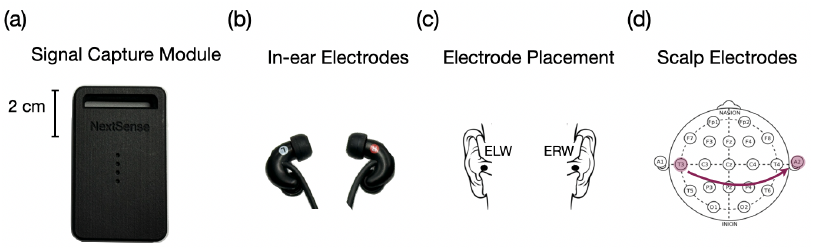
In-ear EEG device configuration. (a) Signal capture module (2 cm scale). (b) In-ear electrodes. (c) Cross-head channel placement (ELW-ERW). (d) Corresponding scalp EEG montage showing T3-A2 channel approximated by the in-ear cross-head configuration.

The final sample included 24 individuals diagnosed with MCI (mean age 74.3 ± 8.6 years 47.9-86.3) and 19 cognitively normal care partner control participants (mean age 68.8±11.4 years (38.7-79.3). Detailed demographic characteristics are presented below in Tables 1 and 2. The sample included 26 females (60.5%) and 17 males (39.5%) with and educational level of 16.9 ± 2.8 years (MCI) and 16.6 ± 1.9 (CN) years with 70.7% holding college degrees or higher. The cohort comprised individuals who were White (74.4%), Black or African American (18.6%), and other racial groups (7.0%).

**TABLE 1.**
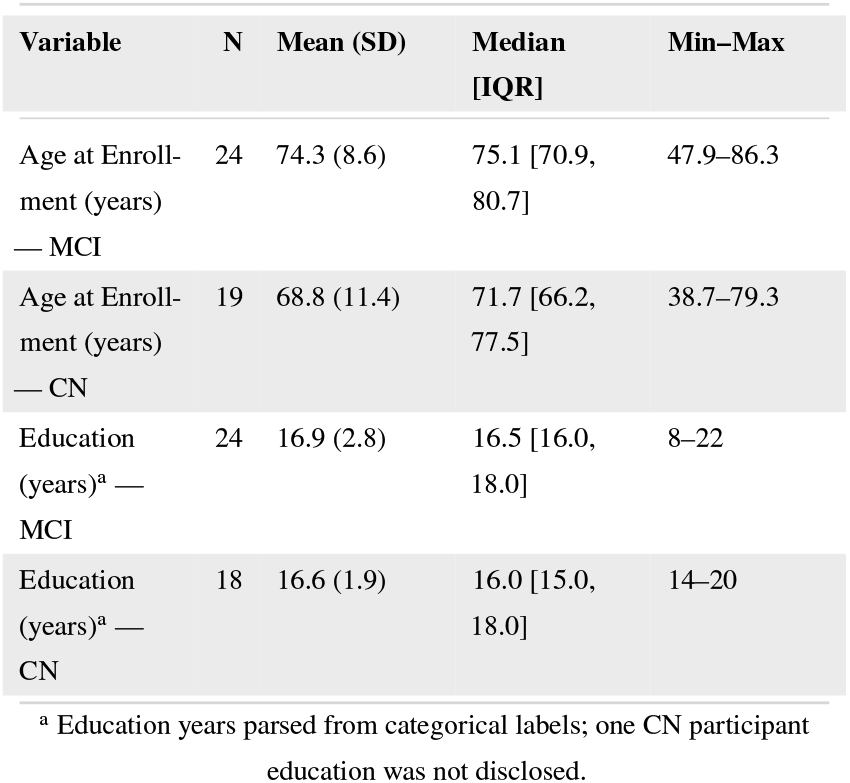
Participant Demographic Information - continuous variables by group.

**TABLE 2.** Participant Demographic Information — categorical variables.

| Variable | Level | N (%) |
| --- | --- | --- |
| Sex | Female | 26 (60.5%) |
|  | Male | 17 (39.5%) |
| Handedness | Right | 38 (88.4%) |
|  | Left | 5 (11.6%) |
| Race | White | 32 (74.4%) |
|  | Black or African American | 8 (18.6%) |
|  | Asian | 1 (2.3%) |
|  | Other Race | 2 (4.7%) |
| Ethnicity | Non-Hispanic | 43 (100.0%) |
| Education (category) | 16 — College graduate | 13 (30.2%) |
|  | 18 — Master's degree | 11 (25.6%) |
|  | 14 — Associate degree | 7 (16.3%) |
|  | 20 — Doctorate | 6 (14.0%) |
|  | 17 | 2 (4.7%) |
|  | 15 | 1 (2.3%) |
|  | 22 | 1 (2.3%) |
|  | 8 | 1 (2.3%) |
|  | Missing | 1 (2.3%) |

Participants underwent longitudinal monitoring as part of a two-phase sleep measurement study that includes at the beginning and end of participation in CEP. Each participant contributed up to seven consecutive nights of at-home sleep recordings during each phase. This multi-night design yielded 361 overnight sessions across all participants (mean 4.44 ± 2.29 nights per participant in each phase).

### 2.2 In-ear EEG Device

Data were recorded using the NextSense Kauai in-ear EEG device with a single cross-head channel configuration (ELW-ERW; Figure 1c). Our preliminary studies with simultaneous scalp and in-ear recordings demonstrated that this channel captures similar patterns to the T3-A2 scalp montage (Figure 1d).

### 2.3 Data Preprocessing

All preprocessing was applied to each recording in its entirety prior to segmentation to keep epoch statistics comparable within a night. Unless stated otherwise, the sampling frequency was *f*_*s*_ = 200 Hz and all steps were implemented in Python 3.9.

### 2.4 Filtering and Normalization

We first removed slow drifts and DC offsets with least-squares linear detrending, then limited the spectrum to sleep-relevant frequencies (0.5–30 Hz) using a zero-phase, 4th-order Butter-worth bandpass applied bidirectionally to avoid phase distortion. Finally, each recording was standardized with z-score normalization (mean 0, SD 1) computed at the recording level so that relative amplitude differences between subsequent epochs within the same night were preserved (Figure 2).

**FIGURE 2.**
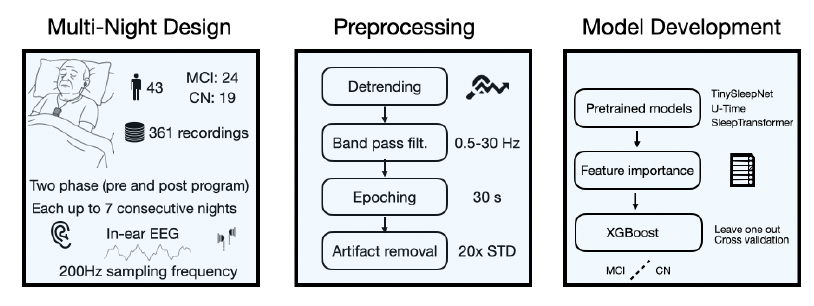
Study pipeline using single-channel in-ear EEG. 43 older adults used in-ear EEG over night leading to 361 sleep recordings. Preprocessing included detrending, bandpass filtering, epoching, and artifact removal. Three pretrained models extracted embeddings, followed by feature selection and XGBoost classification with leave-one-subject-out cross-validation.

#### 2.4.1 Artifact Detection and Segmentation

To maximize usable data while excluding clearly corrupted segments, we retained a 30 s epoch only if it satisfied a set of criteria as follows. Per-epoch rules spanning amplitude, variability, oscillatory behavior, and basic data integrity. Specifically, peak amplitude had to be no more than 20 times the epoch standard deviation; variance had to be at least 10^−4^; and the zero-crossing rate was required to fall within [0.001, 0.95]. All epochs also had to be non-flat (standard deviation ≥ 10^−5^), and no more than 95% of adjacent samples could be identical to guard against frozen traces. This combination rejects obvious artifacts while retaining normal sleep-related variability.

After preprocessing, signals were segmented into non-overlapping 30 s epochs (6000 samples at 200 Hz), consistent with standard sleep scoring practice. The number of epochs per recording varied with duration (mean 708 ± 563 epochs/recording).

We compared four approaches that operate on artifact-free, 30 s epochs and feed into the recording- and subject-level aggregation pipeline described in Section 2.3: three pre-trained deep learning models that return 128-dimensional embeddings per epoch, used strictly in inference mode without fine-tuning, and a traditional baseline based on spectral bandpower. All methods were applied uniformly to the same preprocessed inputs.

#### 2.4.2 AI-driven Embeddings

We extracted 128-dimensional embeddings from each 30 s epoch using three pretrained architectures. TinySleepNet ^[21]^ is a lightweight one-dimensional convolutional neural network composed of convolution–ReLU–pooling blocks with global pooling (∼ 5 × 10^5^ parameters). SleepTransformer ^[22]^ is a transformer with positional encoding and multi-head self-attention whose globally pooled output forms the embedding (∼ 2 × 10^6^ parameters). U-Time^[23]^ is a one-dimensional U-Net (encoder–decoder with skip connections) whose final convolution followed by global pooling yields the embedding (∼ 10^6^ parameters).

As a classical baseline, we computed power spectral density (PSD) per epoch using Welch’s method (Hann windows, 50% overlap), then integrated PSD within standard sleep bands to obtain absolute powers **p**_*k*_ = [*P*_*δ*_, *P*_*θ*_, *P*_*α*_, *P*_*β*_] for *δ* (0.5–4 Hz), *θ* (4–8 Hz), *α* (8–13 Hz), and *β* (13–30 Hz). To reduce inter-subject amplitude variability, we formed relative bandpowers

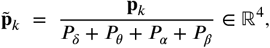

which sum to one and are directly comparable across subjects and nights.

Window-level features (either **f**_*k*_ ∈ ℝ^128^ or 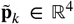) were summarized at the recording level by concatenating the mean, standard deviation, element-wise maximum, and element-wise minimum over all retained epochs, then averaged across nights to form a single subject-level vector (see Section 2.3). This yields 4 × 128 = 512 dimensions for each embedding-based method and 4×4 = 16 dimensions for band-power. Feature selection (e.g., mutual-information ranking) was performed within the classification pipeline to maintain the dynamic of the original feature spaces.

### 2.5 Classification

Within each fold, we first balanced the training set using Synthetic Minority Over-Sampling Technique (SMOTE)^[24]^ applied only to the minority class with five nearest neighbors. We then standardized all features by computing the mean and standard deviation from the training set and applying that transformation to both the training data and the held-out test subject using *x*^′^ = (*x* − *μ*_train_)*/σ*_train_. Next, we performed mutual-information-based feature selection on the training set, retaining the top fifty features for embeddings and all sixteen features for bandpower. Finally, we fit a Gradient Boosting Classifier with a learning rate of 0.3, one hundred estimators, maximum depth of fifteen, and a subsample ratio of 1.0. All preprocessing steps and model fitting were performed exclusively on the training data before being applied to the held-out subject.

We compared two aggregation strategies for best performing model, embeddings: (i) first-night only, in which recording-level features from the first available night were used for each subject and (ii) multi-night averaged, in which recording-level features were computed per night and then averaged across all available nights per subject. For both datasets we used the same subject-level pipeline.

### 2.6 Evaluation Metrics

We used with *n* = 43 subjects, each fold we conducted leave one subject out (LOSO). Essentially we held one subject for testing and used the remaining *n* − 1 subjects for training, ensuring no subject appeared in both sets within a fold. Predictions were concatenated across all folds at the subject level. We report Accuracy, Precision, Recall (Sensitivity), Specificity, F1-score, AUROC, and AUPRC.

#### 2.6.1 Explained Variance

To quantify the contributions of between-subject and within-subject variability to the total variance in sleep EEG embeddings, we performed a variance decomposition analysis across all recording nights and subjects. We calculated three variance components for each embedding dimension, the total variance computed across all recordings from all subjects and nights, the between-subject variance capturing differences between individuals by computing the variance of subject-level means where each subject’s mean was averaged across all their recording nights, and the within-subject variance quantifying night-to-night variability by averaging the variance of recordings within each subject across their nights.

The proportion of total variance explained by each component was calculated as the ratio of that component’s variance to the total variance. These proportions quantify how much of the total variability is attributable to stable inter-individual differences versus transient intra-individual fluctuations, providing insight into the signal-to-noise characteristics of single-night versus multi-night recordings.

#### 2.6.2 Group Separability

To assess how well each model distinguished between MCI and CN groups, we evaluated group separability using predicted probabilities from leave-one-subject-out cross-validation. Cohen’s *d* effect size measured the standardized difference between MCI and CN probability distributions, while Mann-Whitney tests (two-sided) assessed statistical significance. These analyses were performed separately for first-night-only and multi-night-averaged predictions to determine whether aggregating multiple nights improved discriminability. The Mann-Whitney test was chosen for its robustness to non-normal distributions and appropriateness for small sample sizes. First night recordings were selected due to robust availability of data across subjects.

### 2.7 Demographic Adjustment Analysis

To disentangle disease-specific features from demographic influences, we performed both unadjusted and demographically adjusted analyses for the best-performing model. For unadjusted group comparisons, we applied independent samples *t*-tests for normally distributed features (Shapiro-Wilk *p* > 0.05) or Mann-Whitney U tests for non-normally distributed features. We quantified effect sizes using Cohen’s *d*, calculated as the difference between group means 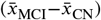 divided by the pooled standard deviation:

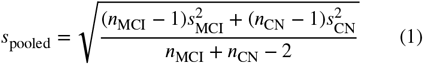

where 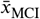 and 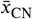 represent the mean feature values for the MCI and CN groups, respectively, and *s*_MCI_ and *s*_CN_ are the corresponding standard deviations.

To assess whether identified features remained significant after accounting for demographic factors, we implemented Analysis of Covariance (ANCOVA) models for each feature, with diagnosis as the primary factor of interest and age (continuous) and sex (binary) as covariates. We computed partial eta squared 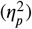 as the effect size measure, calculated as the ratio of the sum of squares attributable to diagnosis to the sum of diagnosis and residual sums of squares.

## 3 RESULTS

### 3.1 Data Quality

After preprocessing and artifact removal, the mean data retention rate was 65.2 ± 34.9%. Longer recordings showed slightly higher retention (*r* = 0.13, *p* = 0.012). Data quality was similar across groups, with a mean retention rate of 62.7% for MCI and 68.4% for CN subjects, despite differences in recording patterns between groups. Figure 3 illustrates these recording characteristics, showing the distribution of mean and standard deviation of epochs across participants, with individual data points color-coded by age.

**FIGURE 3.**
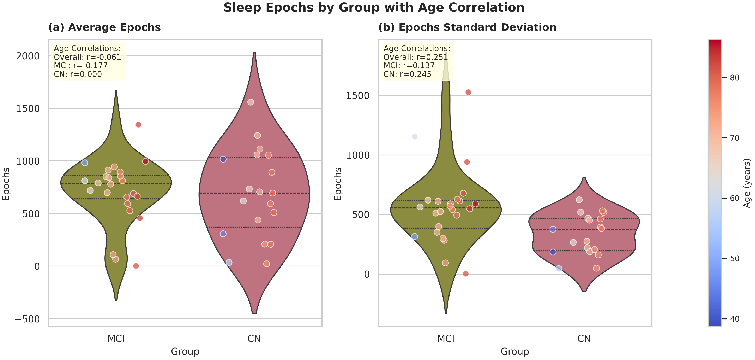
Distribution of sleep epochs by groups with age correlation.(a) Average Epochs and (b) Epochs Standard Deviation for MCI (Mild Cognitive Impairment) and CN (Cognitively Normal) groups. Violin plots show kernel density estimates with quartile lines. Individual points encode participant age by position (left=younger, right=older) and color (blue=younger, red=older). Inset boxes display Pearson correlation coefficients between age and epoch values.

### 3.2 Classification Performance

Using data from all available nights, TinySleepNet and Sleep-Transformer both achieved an overall accuracy of 72.1%, while U-Time reached 60.5% and the bandpower baseline achieved 62.8%. Among the deep models, TinySleepNet yielded the highest sensitivity (79.2%) and F1-score (0.760), indicating strong identification of participants with MCI. Sleep-Transformer provided the best discrimination, with the highest AUROC (0.785) and AUPRC (0.823), and the highest specificity (68.4%). U-Time showed comparable sensitivity (75.0%) and a solid AUPRC (0.791) but lower specificity (42.1%). The bandpower approach delivered moderate performance (AUROC = 0.627, AUPRC = 0.697).

When restricted to first-night data only, performance declined across all methods. TinySleepNet and SleepTransformer maintained the highest accuracy (67.4%), while U-Time and bandpower dropped to 56.6% and 44.2%, respectively. In this scenario TinySleepNet achieved the highest sensitivity (75.0%) and F1-score (0.720), while SleepTransformer showed the best AUROC (0.623), AUPRC (0.627), and specificity (62.7%). The bandpower baseline showed reduced performance with first-night data alone (AUROC = 0.432, AUPRC = 0.541). Table 3 summarizes all quantitative results for both multi-night and single-night conditions, and the corresponding classification performance is illustrated in terms of AUROC and AUPRC (for multi-night) in Figure 4 (a) and (b) respectively.

**TABLE 3.** Performance metrics for MCI classification using sleep EEG.

| Method | Accuracy | Precision | Recall | F1 | AUROC | AUPRC | Sensitivity | Specificity |
| --- | --- | --- | --- | --- | --- | --- | --- | --- |
| All nights |  |  |  |  |  |  |  |  |
| TinySleepNet | 0.721 | 0.731 | 0.792 | 0.760 | 0.774 | 0.799 | 0.792 | 0.632 |
| SleepTransformer | 0.721 | 0.750 | 0.750 | 0.750 | 0.785 | 0.823 | 0.750 | 0.684 |
| U-Time | 0.605 | 0.621 | 0.750 | 0.679 | 0.708 | 0.791 | 0.750 | 0.421 |
| Bandpower | 0.628 | 0.667 | 0.667 | 0.667 | 0.627 | 0.697 | 0.667 | 0.579 |
| First night only |  |  |  |  |  |  |  |  |
| TinySleepNet | 0.674 | 0.692 | 0.750 | 0.720 | 0.614 | 0.609 | 0.750 | 0.579 |
| SleepTransformer | 0.674 | 0.710 | 0.710 | 0.711 | 0.623 | 0.627 | 0.710 | 0.627 |
| U-Time | 0.566 | 0.588 | 0.710 | 0.643 | 0.562 | 0.603 | 0.710 | 0.386 |
| Bandpower | 0.442 | 0.500 | 0.375 | 0.429 | 0.432 | 0.541 | 0.375 | 0.526 |

**FIGURE 4.**
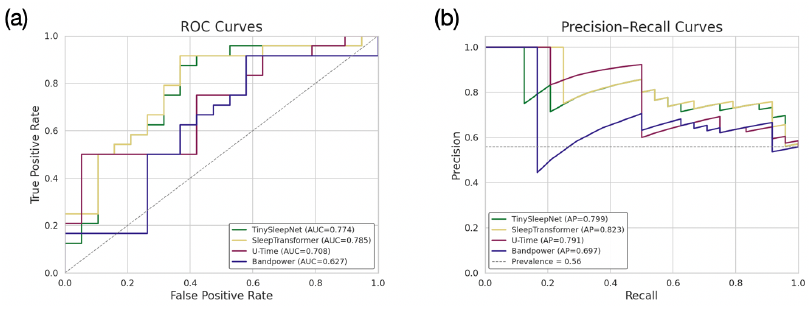
A) ROC and (B) Precision–Recall (PR) curves for MCI classification using embeddings from TinySleepNet, SleepTransfomer, and U-time. Legends report AUC (ROC) and AP (PR); the gray dashed lines indicate chance (ROC) and class prevalence (PR), respectively.

### 3.3 Effect of Multi-Night Averaging

#### 3.3.1 Explained Variance

Between-subject differences explained 41% of total variance in the embeddings, while night-to-night variability within subjects accounted for 91% before averaging. Multi-night averaging reduced feature variance by median factors ranging from 1.29× to 1.60× across embedding methods (approximately 63% to 78% of original variance retained).

#### 3.3.2 Group - Level Separability

Multi-night averaging improved group separability across all methods. TinySleepNet showed Cohen’s *d* increase from 0.65 to 0.96, with Mann-Whitney test *p*-value improving from 0.208 to 0.0023. SleepTransformer demonstrated Cohen’s *d* increase from 0.56 to 1.08, with *p*-value improving from 0.080 to 0.0013. U-Time showed Cohen’s *d* increase from 0.18 to 0.53, with *p*-value improving from 0.599 to 0.021.

### 3.4 Age Correlation Analysis

Feature-age correlation analysis showed that among features selected in each individual fold, 12–20% were significantly correlated with age (*p* < 0.05). The strongest relationships were found in TinySleepNet (12.1%), followed by U-Time (8.0%) and SleepTransformer (6.1%). Age-related features were concentrated among top-ranked features: 29.6% of the ten most important features in TinySleepNet and 19.4% in both SleepTransformer and U-Time exhibited significant correlations with age. In contrast, fewer than 3% of middle-ranked features showed such associations.

### 3.5 Feature Stability Across Folds

Across all leave-one-subject-out cross-validation folds, mutual information-based feature selection identified a total of 162 unique features for TinySleepNet, 172 for SleepTransformer, and 157 for U-Time. Among these selected features, 34 features (20.99%) in TinySleepNet, 32 features (18.60%) in SleepTransformer, and 32 features (20.38%) in U-Time showed significant age correlations (*p* < 0.05) in at least one fold (Figure 5a). This indicates that approximately 19–21% of discriminative features were age-related, while the remaining 79–81% were age-independent. The distribution of correlation coefficients showed consistent patterns across methods, with peaks at *r* = ±0.4 and ranging from −0.5 to +0.5 (Figure 5b).

**FIGURE 5.**
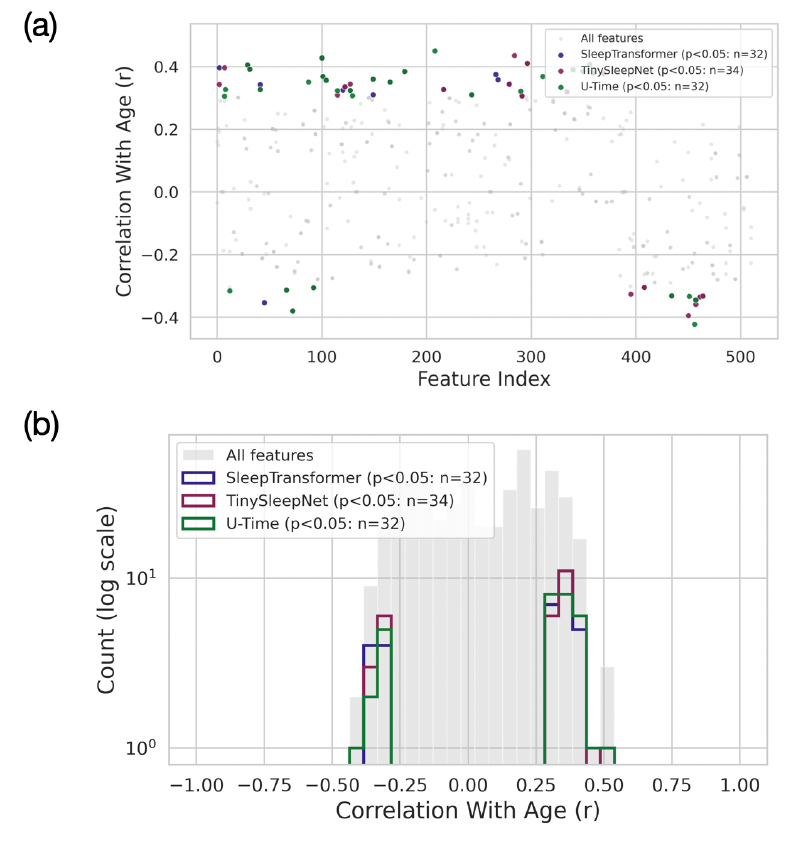
Age-correlated features in the classification pipeline. (a) Features with significant age correlations (*p* < 0.05) selected across all LOSO folds for SleepTransformer (*n* = 32), TinySleepNet (*n* = 34), and U-Time (*n* = 32) shown by feature index. Gray dots represent all features; colored dots indicate significant age correlations for each method. (b) Distribution of correlation coefficients showing consistent patterns across methods with peaks at *r* = ±0.4.

### 3.6 Demographic-Adjusted Feature Analysis

Univariate analysis identified 7 features (1.4%) with significant group differences (*p* < 0.05). After controlling for age and sex using ANCOVA models, 3 of these 7 features (42.9%) maintained significance (*p* < 0.05). Additionally, 4 previously non-significant features became significant after demographic adjustment, revealing disease-specific features that were masked by demographic variance.

## 4 DISCUSSION

This study demonstrates that multi-night, at-home in-ear EEG recordings combined with deep learning models can distinguish older adults with MCI from cognitively normal aging population with clinically relevant accuracy. TinySleepNet achieved the highest F1-score (0.76) and sensitivity (79.2%), while SleepTransformer provided superior discrimination (AUROC = 0.785). Both methods substantially outperformed traditional bandpower features.

### 4.1 Advantages of Deep Learning Embeddings

The superior performance of deep learning embeddings over bandpower features suggests these models capture subtle sleep microarchitectural patterns beyond traditional frequency-band analysis. While bandpower quantifies overall oscillatory activity in predefined frequency ranges, pretrained sleep-staging networks potentially learn hierarchical representations of transient events—such as sleep spindles, K-complexes, and microarousalsthat characterize sleep stage transitions and stability. These transient features are known to be disrupted in early neurodegeneration and may reflect synaptic dysfunction and impaired sleep-dependent memory consolidation processes that precede overt cognitive symptoms ^[25;26]^.

### 4.2 Multi-Night Aggregation Improves Reliability

Aggregating multiple nights of recordings substantially improved classification performance and statistical separability between groups. Accuracy increased from 67.4% to 72.1%, AUROC from 0.614 to 0.774, and effect sizes increased by 48–93% across methods, with previously non-significant group differences becoming clearly significant. This improvement comes from averaging which reduced within-subject night-to-night variability while preserving between-subject differences that reflect stable neurophysiological characteristics. However, the reduction in variability could also be due to measurement noise from environmental factors or transient physiological states.

The variance reduction we observed (median 1.29–1.60× for different models) is smaller than theoretical predictions assuming independent nights, which is expected because consecutive at-home nights share physiological (circadian timing, sleep homeostasis) and environmental factors. Nevertheless, even modest noise reduction was sufficient to enhance both classification metrics and statistical power, demonstrating that single-night assessments underestimate true MCI-CN separability. This finding aligns with repeated-measures theory and emphasizes the value of longitudinal home-based monitoring for reliable feature assessment.

The feasibility of multi-night, in-home recording represents a key practical advantage of in-ear EEG compared to laboratory polysomnography. Participants recorded an average of 4.44 nights at home, a protocol that would be prohibitively expensive and logistically challenging in sleep laboratories. The unobtrusive form factor enables naturalistic sleep monitoring without the first-night adaptation effects that confound laboratory polysomnography, creating a realistic pathway for scalable, naturalistic, and, repeated assessments in community-dwelling older adults.

### 4.3 Disease-Specific Features Beyond Normal Aging

While age contributed to classification, our demographic adjustment analysis demonstrates that MCI-related features extend beyond normal aging. After controlling for age and sex, 42.9% of originally significant features maintained their association with diagnosis, and additional disease-specific markers emerged that were previously masked by demographic variance. This indicates that pathological sleep alterations in MCI represent distinct neurophysiological changes rather than accelerated aging.

The concentration of age-correlated features among top-ranked predictors (29.6% in TinySleepNet, 19.4% in other models) versus middle-ranked features (<3%) suggests that the most discriminative sleep features naturally correlate with age because MCI risk increases with aging. However, the persistence of demographic-independent features after adjustment confirms that our classification relies substantially on pathological processes, likely reflecting amyloid and tau deposition effects on sleep-wake regulatory circuits and glymphatic clearance dysfunction, rather than solely capturing demographic risk factors.

### 4.4 Limitations and Future Works

Next steps should focus on validation in larger, demographically diverse cohorts with longitudinal follow-up to assess predictive validity for cognitive decline. Direct comparison with pTau biomarkers would establish whether sleep-derived features track underlying neuropathology or reflect compensatory mechanisms and downstream consequences. Integration with other digital biomarkers, such as daytime actigraphy during daytime or smartphone-based cognitive assessments (such as P300 auditory oddball task), could also improve classification accuracy of the model.

Beyond monitoring applications, the continuous home-based recording capability of in-ear EEG enables potential therapeutic intervention opportunities. For example, closed-loop auditory stimulation, where sounds are delivered as phase-locked to slow-wave oscillations, has shown promise for enhancing sleep-dependent memory consolidation in laboratory settings ^[27;28]^. Translating these approaches like these to home-based systems which could provide personalized interventions targeting sleep quality in at-risk individuals, potentially slowing progression of cognitive decline. However, such applications require substantial additional development and validation before clinical deployment.

## 5 CONCLUSIONS

Multi-night, single-channel in-ear EEG recorded at home and analyzed with pretrained sleep-staging embeddings distinguished older adults with MCI from cognitively normal peers, reaching an F1-score of 0.76 with TinySleepNet under leave-one-subject-out validation. Aggregating nights (mean 4.44 ± 2.29 nights/participant) substantially improved classification stability, increasing accuracy from 67.4% to 72.1%, AUROC from 0.614 to 0.774, and sensitivity from 62.5% to 75.0%. Demographic adjustment revealed that disease-specific features extend beyond normal aging effects, with 42.9% of significant features maintaining associations after controlling for age and sex. These findings indicate that at-home in-ear EEG provides a feasible route to scalable, longitudinal monitoring of sleep-derived features of mild cognitive impairment risk without requiring in-lab polysomnography, supporting future development as a screening and monitoring tool for early-stage neurodegeneration.

## Abbreviations

MCI: Mild Cognitive Impairment
EEG: Electroencephalography

## 7 ACKNOWLEDGMENTS

YK, AL, and GC acknowledge support from the Alzheimer’s Association, The Michael J. Fox Foundation for Parkinson’s Research, and CurePSP Sleep Contributions to Neurodegeneration Grant Program (SCN-25-1470707). The Charlie and Harriet Shaffer Cognitive Empowerment Program is made possible by a generous gift from the James M. Cox Foundation.

## 8 DATA AVAILABILITY

All data produced in the present study are available upon reasonable request to the authors

